# Anterior Knee Pain Risk Differs Between Male and Female Military Tactical Athletes

**DOI:** 10.1101/2020.09.17.20196741

**Authors:** Neal R. Glaviano, Michelle C. Boling, John J. Fraser

## Abstract

**Background:** Anterior knee pain (AKP) is commonly diagnosed in military members and is a threat to operational readiness. AKP includes a range of conditions, with localized pain around the patella being a frequent location of pain and disability. Mechanical overuse is one suggested etiology for many of these conditions, suggesting occupational demands in the military may influence AKP frequency amongst the service members. Previous research suggests females are at a greater risk for AKP, however, it is unknown how occupation affects AKP risk.

**Study Design:** Epidemiological cohort.

**Level of Evidence:** Level 2.

**Methods:** The Defense Medical Epidemiology Database was queried for the number of individuals with ICD-9 diagnosis codes on their initial encounter from 2006 to 2015. Diagnoses were categorized into anterior or retropatellar pain, patellar instability, knee tendinopathy, as well the sum of the three groups which was defined as AKP. Relative risk (RR) and chi-square statistics were calculated in the assessment of sex and occupational category. Regressions were calculated to determine association between service branch, sex, and AKP across time.

**Results:** From 2006-2018, a total of 151, 263 enlisted and 14,335 officer services members were diagnosed with AKP. The incidence rate was 13.2 in enlisted members and 6.2 in in officers. Females were significantly at greater risk of developing AKP compared to males within both the enlisted (relative risk=1.32) and officer (relative risk=2.01) service members. Differences in risk were also noted across military occupation for both enlisted and officer service members, p<.05.

**Conclusion:** Sex and military occupation were salient factors for AKP risk. Evaluation of training requirements and developing interventions programs across military occupation could serve as a focus for future research aiming to decrease the incidence of chronic knee pain.

## Introduction

Anterior knee pain (AKP) is a common lower extremity injury reported within various populations; adolescents,^1^ physically active individuals,^2^ the general population,^3^ and the military.^4^ AKP is often used as a catch-all term for various types of pathologies that relate to the patella and anterior aspect of the knee. These pathologies often include patellofemoral pain, patellar tendinopathy and patellar instability. While diagnostic criteria vary between the subgroups of AKP, commonalities exist in patient presentation. AKP patients often report long-term recurrent symptoms,^5,6^ decreased subjective function^7^, and limitations related to strenuous activity and sport.^2,8^ Over 91% of patients with AKP will report pain for up to 16 years following initial diagnosis,^9,10^ demonstrating a significant burden on their health-related quality of life.

The military has an alarming incidence and prevalence rate for AKP. In a recent systematic review^3^, the incidence of AKP among military recruits ranged from 9.7-571.4 per 1000 person years.^4,11-16^ Greater incidence rates do exist for females (33 per 1000 person-years) compared to their male counterparts (15 per 1000 person-years).^1,4^ Additionally, the prevalence of AKP has been reported to be as high as 13.5% among naval midshipmen, with 25% of females enrolling at the United States Naval Academy reporting a previous diagnosis of AKP.^4,9^ The high incidence and prevalence rates of AKP among military recruits is concerning when considering the clinical presentation and the chronicity of symptoms. AKP presents with knee pain during weight-bearing tasks, such as walking, running, jumping, squatting, lunging, and stair ambulation.^17^ Individuals experiencing pain during these tasks limit their ability to perform activities of daily living, recreational exercise, and perform their occupation,^18,19^ which could be very detrimental to individuals serving in the military. The functional limitations due to AKP account for a larger proportion of medical discharge from the military compared to other lower extremity injuries.^11,15^

Because AKP is a common condition among military recruits and is exacerbated during functional tasks required for members of the military to complete, it is important to understand if the incidence of this condition differs across diverse military occupations. This is important since disparity in physical exposure to varying load carriage requirements, physical training, kneeling, and jumping may exist between military occupations. Furthermore, potential sex-related factors of AKP within military occupation (with the assumption of similar occupational exposure) is currently unknown. The identification of AKP risk across sex and military occupation would allow for greater precision in resource allocation and development of more ecologically valid treatment approaches, targeted interventions groups with the greatest need, and improvement of military readiness. Therefore, the purpose of this study is to assess the risk of AKP in military servicemen and women across multiple military occupations.

## METHODS

A population-based epidemiological retrospective cohort study of all service members in the US Armed Forces was performed assessing risk of sex and military occupation on the outcome of AKP incidence from 2006-2015. The Defense Medical Epidemiological Database [(DMED), Defense Health Agency, Falls Church, VA, https://bit.ly/DHADMED] was utilized to identify relevant healthcare encounters. This database provides aggregated data for International Classification of Diseases, Ninth Revision (ICD-9) codes and de-identified patient characteristics, including sex, categories of military occupations, and branch of service for all active duty and reserve military service members. The database is HIPAA compliant, does not include any personal identifiable or personal health information, and has been used previously for epidemiological study of lower extremity injury in the military.^20,21^ This study was approved as non–human-subjects research by the Institutional Review Board at the Naval Health Research Center (NHRC.2020.0203-NHSR).

Since AKP is characterized by a diverse set of potential diagnoses, the database was queried for the number of distinct patients with a primary diagnosis of antero- and retropatellar pain (717.7, Chondromalacia patella; 726.65 prepatellar bursitis), patella instability (ICD-9 codes 718.36, patella subluxation; 836.3, patellar dislocation), and knee tendinopathy [726.60 quadricep tendinopathy (enthesopathy of knee, unspecified); 726.64, patellar tendinopathy], on the initial medical encounter from 2006–2015. Patients with repeat visits for the same diagnosis were only counted once in all analyses.

Calculations of cumulative incidence of patients diagnosed with AKP were conducted for male and female military members, enlisted and officers, in each service branch (Army, Navy, Marine Corps, and Air Force) and occupational category. Relative risk (RR) point estimates and 95% confidence intervals (CIs), risk difference point estimates, attributable risk (AR), number needed to harm (NNH), and chi-square statistics were calculated in the assessment of sex and occupation category, using the group with the lowest rate as the reference group. Linear regression was performed to evaluate branch, rank, sex, and year on the incidence of AKP using R Version 3.5.1 (The R Foundation for Statistical Computing, Vienna, Austria). The level of significance was *p* ≤ 0.05 for all analyses. RR point estimates were considered statistically significant if CIs did not cross the 1.00 threshold. All calculations were performed using Microsoft Excel for Mac 2016 (Microsoft Corp., Redmond, WA) and a custom epidemiological calculator spreadsheet.^22^

## RESULTS

From 2006 to 2015, 151,263 enlisted service members were diagnosed with AKP; 60,670 diagnosed with anterior or retro-patellar pain, 11,956 diagnosed with patellar instability, and 78,637 diagnosed with patellar tendinopathy. A total of 14,335 military officers were diagnosed with AKP; 10,105 diagnosed with anterior or retro-patellar pain, 1,014 diagnosed with patellar instability, and 3,216 diagnosed with patellar tendinopathy. The total incidence rate of AKP in enlisted service members was 13.2 per 1,000 person-years, while military officers was 6.2 per 1,000 person-years. There was no statistically significant change in AKP risk between 2006-2015 (*t*=.650, *p*=.517).

### Sex

Numbers of AKP cases and incidence between sex is reported for enlisted services members in Table 1 and officers in Table 2. Enlisted females had an incidence rate of 16.7 per 1000 person-years compared to the enlisted male service members with an incidence rate of 12.7 per 1000 person-years (RR: 1.32, 95%CI: 1.30-1.34, p<0.001) across all AKP diagnoses. Females enlisted in the Army had the greatest risk of AKP, 20.9 per 1000 person-years. Female enlisted service members also had significantly greater incidence risk for tendinopathy (RR: 1.28, 95%CI: 1.26-1.31, p<0.001), instability (RR: 1.56, 95%CI: 1.49-1.63, p<0.001), and anterior- and retro-patellar pain (RR: 1.32, 95%CI: 1.30-1.35, p<0.001). (Table 3) Female officers had an incidence rate of 10.7 per 1,000 person-years compared to male officers at an incidence rate of 5.3 per 1000 person-years (RR: 2.01, 95%CI: 1.94-2.09). Female officers had the greatest relative risk of experiencing tendinopathy (RR: 4.75, 95%CI:4.43-5.09) when compared to male officers.

**Table 1.** Numbers and incidence of anterior knee pain among enlisted members of the US Armed Forces

|  | Males | Females | Total | Males | Females | Total |
| --- | --- | --- | --- | --- | --- | --- |
| Anterior- and Retropatellar pain (n=60670) |  |  | Incidence Rate<br>(per 1000 person-years) |  |  |  |
| Army | 23052 | 4986 | 28038 | 6.1 | 8.7 | 6.4 |
| Navy | 6958 | 1814 | 8772 | 3.0 | 4.1 | 3.2 |
| Air Force | 11165 | 3571 | 14736 | 5.3 | 7.0 | 5.6 |
| Marines | 8480 | 644 | 9124 | 5.3 | 5.5 | 5.3 |
| Total | 49655 | 11015 | 60670 | 5.1 | 6.7 | 5.3 |
| Patellar Instability (n=11956) |  |  | Incidence Rate<br>(per 1000 person-years) |  |  |  |
| Army | 3515 | 812 | 4327 | 0.9 | 1.4 | 1.0 |
| Navy | 1881 | 567 | 2448 | 0.8 | 1.3 | 0.9 |
| Air Force | 2293 | 899 | 3192 | 1.1 | 1.8 | 1.2 |
| Marines | 1791 | 198 | 1989 | 1.1 | 1.7 | 1.2 |
| Total | 9480 | 2476 | 11956 | 1.0 | 1.5 | 1.0 |
| Knee Tendinopathy (n=78637) |  |  | Incidence Rate<br>(per 1000 person-years) |  |  |  |
| Army | 31413 | 6206 | 37619 | 8.3 | 10.8 | 8.6 |
| Navy | 9457 | 2690 | 12147 | 4.1 | 6.1 | 4.5 |
| Air Force | 14608 | 3902 | 18510 | 6.9 | 7.7 | 7.1 |
| Marines | 9251 | 1110 | 10361 | 5.8 | 9.4 | 6.0 |
| Total | 64729 | 13908 | 78637 | 6.6 | 8.5 | 6.9 |
| Total Anterior Knee Pain (n=151263) |  |  | Incidence Rate<br>(per 1000 person-years) |  |  |  |
| Army | 57980 | 12004 | 69984 | 15.3 | 20.9 | 16.1 |
| Navy | 18296 | 5071 | 23367 | 8.0 | 11.5 | 8.6 |
| Air Force | 28066 | 8372 | 36438 | 13.3 | 16.5 | 13.9 |
| Marines | 19522 | 1952 | 21474 | 12.2 | 16.6 | 12.5 |
| Total | 123864 | 27399 | 151263 | 12.7 | 16.7 | 13.2 |

**Table 2.** Numbers and incidence of anterior knee pain among officers of the US Armed Forces

|  | Males | Females | Total | Males | Females | Total |
| --- | --- | --- | --- | --- | --- | --- |
| Anterior- and Retropatellar pain (n=10105) |  |  | Incidence Rate<br>(per 1000 person-years) |  |  |  |
| Army | 4023 | 1052 | 5075 | 5.2 | 7.1 | 5.5 |
| Navy | 1300 | 312 | 1612 | 2.9 | 3.7 | 3.1 |
| Air Force | 2010 | 709 | 2719 | 3.8 | 5.8 | 4.2 |
| Marines | 626 | 73 | 699 | 3.2 | 5.7 | 3.3 |
| Total | 7959 | 2146 | 10105 | 4.1 | 5.9 | 4.4 |
| Patellar Instability (n=1014) |  |  | Incidence Rate (per 1000<br>person-years) |  |  |  |
| Army | 293 | 102 | 395 | 0.4 | 0.7 | 0.4 |
| Navy | 141 | 50 | 191 | 0.3 | 0.6 | 0.4 |
| Air Force | 237 | 114 | 351 | 0.4 | 0.9 | 0.5 |
| Marines | 70 | 7 | 77 | 0.4 | 0.5 | 0.4 |
| Total | 741 | 273 | 1014 | 0.4 | 0.7 | 0.4 |
| Knee Tendinopathy (n=3216) |  |  | Incidence Rate (per 1000<br>person-years) |  |  |  |
| Army | 848 | 703 | 1551 | 1.1 | 4.8 | 1.7 |
| Navy | 272 | 259 | 531 | 0.6 | 3.1 | 1.0 |
| Air Force | 420 | 498 | 918 | 0.8 | 4.1 | 1.4 |
| Marines | 158 | 58 | 216 | 0.8 | 4.5 | 1.0 |
| Total | 1698 | 1518 | 3216 | 0.9 | 4.1 | 1.4 |
| Total Anterior Knee Pain (n=14335) |  |  | Incidence Rate (per 1000<br>person-years) |  |  |  |
| Army | 5164 | 1857 | 7021 | 6.6 | 12.6 | 7.6 |
| Navy | 1713 | 621 | 2334 | 3.9 | 7.4 | 4.4 |
| Air Force | 2667 | 1321 | 3988 | 5.0 | 10.8 | 6.1 |
| Marines | 854 | 138 | 992 | 4.4 | 10.8 | 4.8 |
| Total | 10398 | 3937 | 14335 | 5.3 | 10.7 | 6.2 |

**Table 3.** Assessment of risk of anterior knee pain (AKP) by sex in members of US Armed Forces, 2006-2018

| Enlisted Specialty | Artillery/<br>Gunnery | Aviation | Engineers | Maintenance | Administration,<br>Intelligence, &<br>Communication | Logistics | Maritime/<br>Naval<br>Specialties | Enlisted Total |  |  |  |
| --- | --- | --- | --- | --- | --- | --- | --- | --- | --- | --- | --- |
|  |  |  |  |  |  |  |  | Tendinopathy | Instability | Antero- &<br>Retro-<br>patellar | All AKP<br>Diagnoses |
| RR<br>(95% CI) | 1.33<br>(1.16-1.52) | 1.61<br>(1.33-1.95) | 0.42<br>(0.32-0.56) | 1.27<br>(1.23-1.31) | 1.25<br>(1.23-1.28) | 1.24<br>(1.20-1.29) | 1.45<br>(1.27-1.66) | 1.28<br>(1.26-1.31) | 1.56<br>(1.49-1.63) | 1.32<br>(1.30-1.35) | 1.32<br>(1.30-1.34) |
| Risk<br>difference<br>(per 1000<br>person years) | 4.5 | 5.7 | -21.9 | 3.1 | 3.3 | 3.4 | 3.8 | 1.9 | 0.5 | 1.6 | 4.0 |
| AR % | 24.8 | 37.9 | -136.4 | 21.2 | 20.2 | 19.5 | 31.2 | 22 | 35.8 | 24.4 | 24.2 |
| p-value | **<.001** | **<.001** | **<.001** | **<.001** | **<.001** | **<.001** | **<.001** | **<.001** | **<.001** | **<.001** | **<.001** |
| NNH | 224 | 175 | -46 | 323 | 299 | 295 | 264 | 536 | 1849 | 609 | 247 |
| Officer Specialty | Ground /<br>Naval<br>Gunfire | Aviation | Engineering<br>&<br>Maintenance | Administration | Operations &<br>Intelligence | Logistics | Services | Officer Total |  |  |  |
|  |  |  |  |  |  |  |  | Tendinopathy | Instability | Antero- &<br>Retro-<br>patellar | All AKP<br>Diagnoses |
| RR<br>(95% CI) | 0.96<br>(0.79-1.17) | 1.31<br>(1.11-1.55) | 1.28<br>(1.16-1.41) | 1.23<br>(1.10-1.36) | 1.01<br>(0.85-1.19) | 1.31<br>(1.19-1.45) | 1.24<br>(1.16-1.32) | 4.75<br>(4.43-5.09) | 1.96<br>(1.70-2.25) | 1.43<br>(1.37-1.50) | 2.01<br>(1.94-2.09) |
| Risk<br>difference<br>(per 1000<br>person years) | -0.3 | 1.7 | 2.8 | 2.1 | 0.0 | 3.5 | 1.8 | 3.3 | 0.4 | 1.8 | 5.4 |
| AR % | -3.8 | 23.6 | 21.8 | 18.4 | 0.9 | 23.9 | 19.1 | 78.9 | 48.9 | 30.2 | 50.3 |
| p-value | .71 | **.001** | **<.001** | **<.001** | .91 | **<.001** | **<.001** | **<.001** | **<.001** | **<.001** | **<.001** |
| NNH | -3322 | 581 | 364 | 475 | 24867 | 287 | 548 | 306 | 2746 | 566 | 185 |

Female service members referenced to male members. RR, Relative Risk; CI, confidence interval; AR, Attributable Risk; NNH, Number Needed to Harm

### Branch and Rank

The comparison of incidence rate across the individual military services between 2006-2015 are reported in Figure 1. Service members in the Army were at a greater risk of AKP, with enlisted service members having a rate of 16.1 per 1000 person-years and officers at a rate of 7.7 per 1000 person-years (*t*=5.272, *p*<.001). Those in the Navy were at a significantly lesser risk of AKP, enlisted at a rate of 8.6 per 1000 person-years and officers at a rate of 4.4 per 1000 person years (*t*=-7.893, *p*<.001). Military officers were at a lower risk of AKP compared to their enlisted counterparts (*t*=-20.12, *p*<.001).

**Figure 1:**
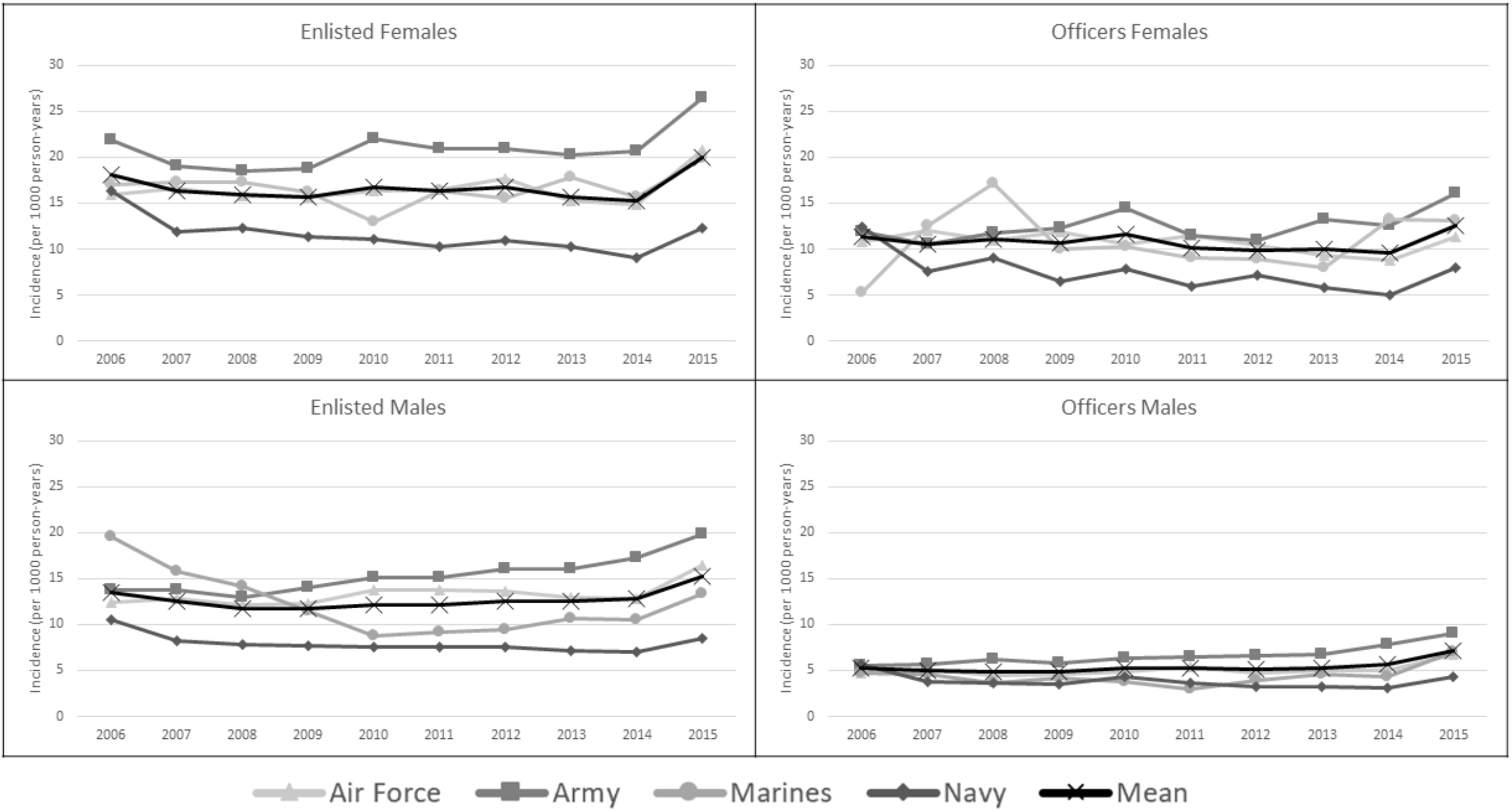
Risk of AKP in Enlisted and Officer Service Members.

### Occupation

Risk of AKP by occupation for both enlisted and officer specialty is reported in Table 4. Enlisted infantry and naval gunfire officers were used as the reference group for comparisons. Significant differences were identified in six enlisted specialty, Mechanized/Armor, Artillery/Gunnery, Engineers, Maintenance, Administration/Intelligence/Communication, and Logistics. The greatest risk of enlisted servicemembers were identified in Logistics (RR: 1.38, 95%CIs: 1.35-1.42) and Administrative/ Intelligence/Communication occupations (RR: 1.33, 95%CIs: 1.30-1.46) compared to enlisted infantry service members. When comparing occupation amongst officers, Administration, Engineering/Maintenance, Operations/Intelligence, and Logistics were statistically significant. The occupations with the greatest risk in officers were Logistics (RR: 1.42, 95%CIs: 1.34-1.50) and Administration (RR: 1.35, 95%CIs: 1.28-1.43) compared to the naval gunfire officers.

**Table 4.** Assessment of risk of anterior knee pain (AKP) by occupation of US Armed Forces, 2006-2015.

| **Enlisted Specialty\*** | Special<br>Operation<br>Forces | Mechanized/<br>Armor | Artillery/<br>Gunnery | Aviation | Engineers | Maintenance | Administration,<br>Intelligence, &<br>Communication | Logistics | Maritime/<br>Naval<br>Specialties |
| --- | --- | --- | --- | --- | --- | --- | --- | --- | --- |
| RR (95% CI) | 0.69<br>(0.64-0.76) | 1.08<br>(1.02-1.15) | 1.31<br>(1.26-1.36) | 0.93<br>(0.87-0.99) | 1.21<br>(1.15-1.27) | 1.12<br>(1.10-1.15) | 1.33<br>(1.30-1.36) | 1.38<br>(1.35-1.42) | 0.86<br>(0.81-0.92) |
| Risk difference (per<br>1000 person years) | -3.2 | 0.8 | 3.2 | -0.8 | 2.2 | 1.3 | 3.5 | 4.0 | -1.4 |
| AR % | -43.9 | 7.5 | 23.6 | -7.9 | 17.5 | 11.0 | 25.0 | 27.8 | -15.8 |
| p-value | **<.001** | **.01** | **<.001** | **.02** | **<.001** | **<.001** | **<.001** | **<.001** | **<.001** |
| NNH | -312 | 1180 | 308 | -1308 | 450 | 768 | 286 | 248 | -698 |
| **Officer Specialty†** | Aviation | Engineering<br>&<br>Maintenance | Administration | Operations &<br>Intelligence | Logistics | Services |  |  |  |
| RR (95% CI) | 0.68<br>(0.65-0.72) | 1.23<br>(1.17-1.30) | 1.35<br>(1.28-1.43) | 1.17<br>(1.11-1.23) | 1.42<br>(1.34-1.50) | 1.00<br>(0.96-1.05) |  |  |  |
| Risk difference (per<br>1000 person years) | -2.6 | 1.9 | 2.9 | 1.4 | 3.5 | 0.0 |  |  |  |
| AR % | -46.5 | 18.8 | 26.1 | 14.4 | 29.6 | 0.2 |  |  |  |
| p-value | **<.001** | **<.001** | **<.001** | **<.001** | **<.001** | .95 |  |  |  |
| NNH | -381 | 520 | 343 | 718 | 287 | 72979 |  |  |  |

\* Contrasted to Enlisted Infantry. † Referenced to Ground and Naval Gunfire Officers. RR, Relative Risk; CI, confidence interval; AR, Attributable Risk; NNH, Number Needed to Harm

## DISCUSSION

The primary finding of this study were sex, rank, and military occupation were salient factors in the risk of AKP. Enlisted females were at 1.32-fold increased risk of AKP compared to enlisted males, while female officers were at a 2.01-fold increase for risk of AKP compared to male officers.

Additionally, females were at greater risk of tendinopathy, instability, and anterior- or retro-patellar pain, with female officers having the greatest relative risk for patellar tendinopathy by 4.75 (95% CI: 4.43-5.09). Finally, military service members who work in logistics have an increased risk of AKP, as shown in both the enlisted (RR:1.38, 95%CI: 1.35-1.42) and officer (RR: 1.42, 95%CI: 1.34-1.50) ranks.

We found the total incidence for AKP in the active duty military to be 13.2 and 6.2 per 1000 person-years in enlisted and officer ranks, respectively. The incidence from the current study are greater than previously established incidence for chondromalacia patellae in the military (4.32 cases per 1000 person-years).^23^ These findings are consistent with previous patellofemoral pain incidence data within the military setting, which ranges between 9.7-571.4 cases per 1000 person-years.^12-16^ Patellofemoral pain and chondromalacia patellae are two specific conditions that can fall within the umbrella term of AKP, which does warrant some caution when comparing incidence rates across studies.

The definition of AKP utilized in this investigation included three subgroups, antero- or retro-patellar pain, patellar instability, and tendinopathy, which provides a unique overview of various forms of knee pain that originate around the patella. Knee tendinopathy had the greatest incidence in enlisted servicemembers of the three subgroups, accounting for 6.9 cases per 1000 person-years. Owens et al.,^24^ identified 584 cases of patellar tendinopathy from 80,106 active duty servicemembers over three separate single-year time periods (2001, 2004 and 2007), but did not report incidence rate in person-years. When comparing the incidence rate of patellar tendinopathy in our military cohort with the general population (1.6 cases per 1000 person-years)^25^, there is a much greater incidence of this condition in the military.^24^ Prevention programs need to be prioritized, as reoccurrence rate ranged between 21-27%^26,27^, resulting in a recurrent decrease in military readiness.

### Sex

Females in both the officer and enlisted ranks had an incidence rate of 10.7 and 16.7 (per 1000 person-years) with an increased relative risk of AKP risk by 2.01 and 1.32 as compared to their male counterparts, respectively. The increased risk of AKP in females is supported by previous patellofemoral pain evidence, with females being 2.23 times more likely to experience this chronic knee condition.^4^ The average incidence rate of chondromalacia has also been previously reported as 4.32 per 1000 person-years across all branches of the military, with females having an adjusted incidence rate ratio of 1.50 (95% CI: 1.47-1.54).^23^ When evaluating the AKP subcategory of antero-retro-patellar pain, which includes the chondromalacia patella ICD-9 code, we had similar incidence rates of 5.3 and 4.4 cases per 1000 person-years in enlisted and officer ranks, respectively. While our data does show similar incidence rates, it should be noted the pathological conditions included in our definition of AKP are broader than the specific diagnosis of patellofemoral pain^4,12-14,16^ or chondromalacia patella.^23^

Females did present with a greater incidence rate for patellar tendinopathies, for both enlisted and officers, compared to their male counterparts. While there is limited evidence evaluating patellar tendinopathy incidence rate across sex, prevalence data suggests males are at a greater risk.^28^ Our findings conflict with previous research, which have speculated that males are at greater risk for developing patellar tendinopathy due to athletic demands and weekly participation in sports.^29,30^ It may be possible that patellar tendinopathy incidence risk may be impacted by intrinsic factors, such as occupation and lower extremity function such as strength. Increased demands in the military could explain why females had a greater incidence rate in the current study, as a linear relationship exists between patellar tendinopathy and training volume.^31^ Future evidence should objectively assess the mechanical overload on the tendon during military training to better understand the higher incidence rate in females and provide direction for appropriate interventions.

### Rank and Branch

Findings from this current study identified that enlisted service members were at a greater risk for AKP than officers. Our cohort of enlisted personnel included both junior and senior enlisted members, but previous research has identified that junior enlisted and senior personnel distinctly have the highest incidence rate of chondromalacia patellae, 3.75, 95% CI:3.57-3.93 and 2.29, 95% CI: 2.20-2.39, respectively compared to military officers.^23^ Additionally, our data compliments previous evidence^23,24,32^ that those in land-based service branches (Army and Marines) are at greater risk for knee injuries than non-land-based service branches (Air Force and Navy). The requirements of both enlisted and land-based service branches suggest that increased activity and training demands continue to increase the risk of developing knee pain. While modification of military requirements is not plausible for all branches, implementation of intervention programs that target those high-risk groups are warranted.

### Occupation

Both enlisted and officer service members had the greatest risk of AKP when employed in logistics, with increased incidence also present in administration, intelligence and communication, engineers, maintainers, and artillery occupations as well. The lowest risk was identified in enlisted special operator and maritime/naval occupations and in the aviation community. Our data agrees with previous evidence evaluating chondromalacia patellae,^23^ patellar tendinopathy,^24^ and other forms of knee injuries among various occupations.^32^ These findings are interesting, as overuse and increased demands are suggested to be the etiology of AKP, and those greater demand occupations reported lower incidence rates. It could be theorized that those in greater demanding occupations, such as infantry, are accustom to the occupational demands. However, it is also plausible that those in greater demanding occupations might not have the same availability to medical care or are less likely to actively seek care due to psychosocial determinants unique to the military.^33^ Objective assessment of the required training load across military occupations would be needed to better understand its role on the incidence rate of AKP.

#### Clinical and Research Implications

This is one of the first studies to evaluate AKP in the active military across branches and occupations. Most of the established incidence rates have been reported in military recruits and cadets enrolling in a 6-week to 25-week training program, which may explain the wide range incidence rates in the previous studies.^12-16^ Increased physical demands and over-use likely contribute to the development of AKP,^34^ as incidence rate of AKP is greatest in the early phases of training and decreased after the first month of training.^15^ AKP does account for the largest cause of medical discharge from military recruits,^11^ however, there is limited evidence evaluating AKP in services members beyond their initial training. Almost 3% of all military service members seek rehabilitative care for knee pain,^15^ supporting our findings that AKP still presents a large concern in the military. Identifying those at greater risk across branches and occupations provides future direction into developing training programs to reduce occupational attrition and improve military readiness. Colocation of uniformed and civilian rehabilitation specialists, which include both physical therapists and athletic trainers, in proximity to the operational forces is warranted in-garrison and while deployed.^33^ When complemented with sports medicine physicians, the diverse skills of the cohesive interdisciplinary musculoskeletal team provide a capacity to perform preventative screening and interventions to address identified risk factors, as well as timely comprehensive treatment following AKP symptom onset.

AKP presents with a heterogeneous presentation of symptoms and impairments, stemming from increased demands or stress on the patellofemoral joint. Those in the Army have the greatest risk of experiencing AKP, which can be troublesome with the recent advancements in the Army’s physical fitness test. The new Army Combat Fitness Test requires a minimum deadlift of 140 pounds, a sprint/drag/carry task which includes carrying an external load and a 2-mile run. These tasks replicate combat conditions but are also tasks that are often pain provoking for individuals experiencing AKP. Additionally, the minimum requirements are age- and sex-neutral, which presents with potential challenges for females who are already at a greater risk for AKP and the influence of age on incidence rates of specific forms of AKP, such as patellofemoral pain or patellar tendinopathy.^23,24^ The inclusion of these tasks will increase the demands of physical readiness of those in the Army to train and complete activities that require an increased external load. Injury surveillance and future epidemiological studies should evaluate incidence rates of AKP conditions after the implementation of the Army Combat Fitness Test.

In athletic populations patellar tendinopathy has been suggested to be related to decreased knee and hip range of motion during functional tasks.^34^ While the physical requirements across military occupations are unknown, evaluating the daily demands might provide insight into the high rate of this condition. Nonetheless, implementation of load management and eccentric training may be warranted as a treatment program for those servicemembers presenting with patellar tendinopathies.^35^ Additionally incorporating quadriceps strengthening programs should implemented across branches, with a focus on land-based groups, to minimize the risk of patellofemoral pain.^36^ These implementation programs should be evaluated for their efficacy at decreasing AKP risk in the military, as they have been shown to be beneficial in both clinical and research settings.^35,37,38^

#### Strengths and Limitations

Using the DMED allowed for a large, population-based analysis of all service members diagnosed with AKP in each branch of military service. The ability to exclude repeat encounters resulted in an estimate of incidence and gold standard epidemiological measures of risk. Further, the stratification by sex addressed a timely military issue of public health importance given that the full integration of women is currently under way. This is the first study to evaluate AKP risk amongst military occupation across all branches of service.

There are limitations to this study. There are a wide range of ICD-9 codes for AKP diagnosis, with a lack of agreement by healthcare professionals. The use of multiple ICD-9 codes for the three subdivisions of AKP minimize the potential bias by providing a range of common codes used in the diagnosis, however other codes may be applicable among healthcare providers. The variability in diagnosis would result in either overestimation or underestimation of AKP across the military branches. Evaluation of ICD-9 selection for chronic knee pain conditions would provide insight into the included codes for this study and future epidemiological studies. Recent clinical practice guidelines for patellofemoral pain^39^ have provided clinician recommendations of both ICD-10 codes and International Classifications of Functioning, Disability, and Health guidelines outlined by the World Health Organization. These recommendations should provide more structure when utilizing large database studies for challenging pathologies that often include a wide range of conditions. Additionally, the data presented in this study only corresponds to active military service members who sought care. Disparity in care-seeking behaviors may introduce a utilization bias in this analysis. Future assessment within occupational groups is warranted using direct survey. The incidence rate of National Guard or Reserve servicemembers is currently unknown. Military service members have demonstrated decreased care-seeking behaviors in other healthy conditions, related to self-reliance, emotional control, the stigma of weakness, or negative beliefs from co-workers.^40,41^ Decreased care-seeking behaviors also exist in community populations experiencing chronic knee pain.^42^ These barriers should be considered since the data included in this current study is from a military population experiencing a chronic knee pain condition.

## CONCLUSION

The average incidence of AKP amongst active duty services members was 13.2 per 1000 person-years in enlisted and 6.2 per 1000 person-years in officers from 2006-2015. Sex and occupation were salient risk factors for AKP, with females and occupations in logistics, administration, intelligence and engineering being at greater risk. Further research is warranted to identify modifiable risk factors and effective treatment plans for these groups at greater risk for AKP.

## Data Availability

The data that support the findings of this study are available from the corresponding author upon reasonable request.

---

**Appendix A.**
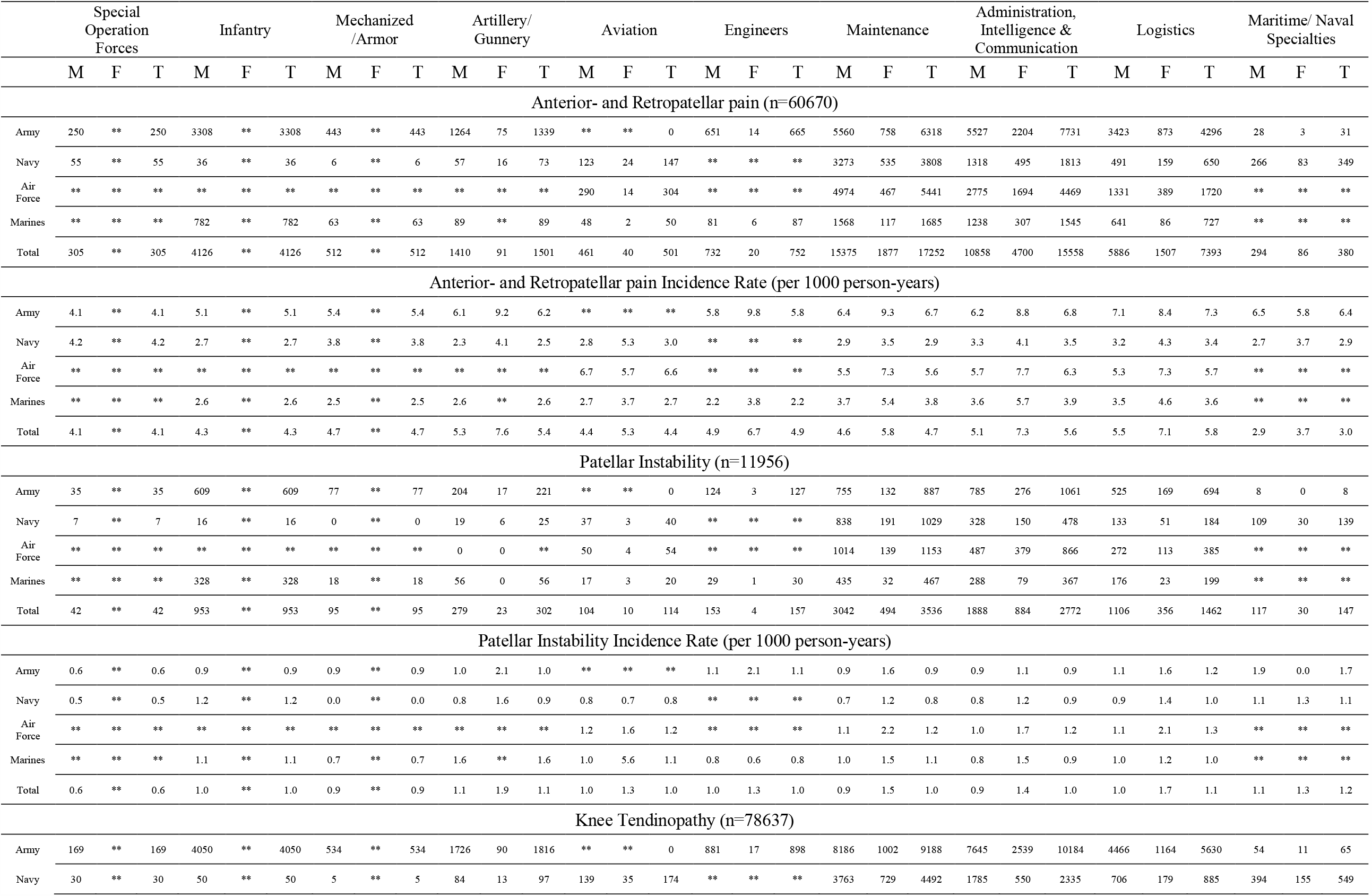
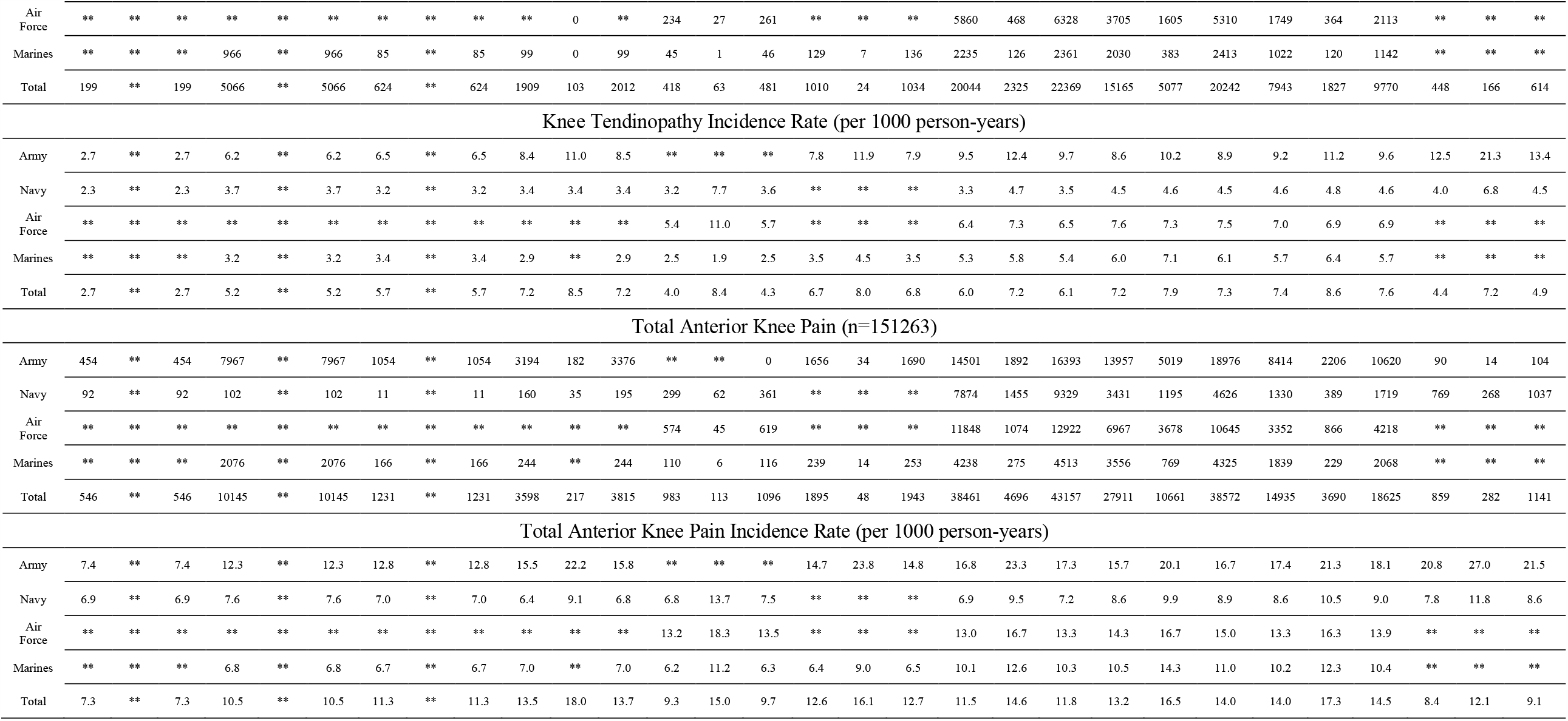
Numbers and incidence of anterior knee pain among enlisted members in the US Armed Forces across occupation.

|  | Special<br>Operation<br>Forces |  |  | Infantry |  |  | Mechanized<br>/Armor |  |  | Artillery/<br>Gunnery |  |  | Aviation |  |  | Engineers |  |  | Maintenance |  |  | Administration,<br>Intelligence &<br>Communication |  |  | Logistics |  |  | Maritime/ Naval<br>Specialties |  |  |
| --- | --- | --- | --- | --- | --- | --- | --- | --- | --- | --- | --- | --- | --- | --- | --- | --- | --- | --- | --- | --- | --- | --- | --- | --- | --- | --- | --- | --- | --- | --- |
|  | M | F | T | M | F | T | M | F | T | M | F | T | M | F | T | M | F | T | M | F | T | M | F | T | M | F | T | M | F | T |
| Anterior- and Retropatellar pain (n=60670) |  |  |  |  |  |  |  |  |  |  |  |  |  |  |  |  |  |  |  |  |  |  |  |  |  |  |  |  |  |  |
| Army | 250 | ** | 250 | 3308 | ** | 3308 | 443 | ** | 443 | 1264 | 75 | 1339 | ** | ** | 0 | 651 | 14 | 665 | 5560 | 758 | 6318 | 5527 | 2204 | 7731 | 3423 | 873 | 4296 | 28 | 3 | 31 |
| Navy | 55 | ** | 55 | 36 | ** | 36 | 6 | ** | 6 | 57 | 16 | 73 | 123 | 24 | 147 | ** | ** | ** | 3273 | 535 | 3808 | 1318 | 495 | 1813 | 491 | 159 | 650 | 266 | 83 | 349 |
| Air Force | ** | ** | ** | ** | ** | ** | ** | ** | ** | ** | ** | ** | 290 | 14 | 304 | ** | ** | ** | 4974 | 467 | 5441 | 2775 | 1694 | 4469 | 1331 | 389 | 1720 | ** | ** | ** |
| Marines | ** | ** | ** | 782 | ** | 782 | 63 | ** | 63 | 89 | ** | 89 | 48 | 2 | 50 | 81 | 6 | 87 | 1568 | 117 | 1685 | 1238 | 307 | 1545 | 641 | 86 | 727 | ** | ** | ** |
| Total | 305 | ** | 305 | 4126 | ** | 4126 | 512 | ** | 512 | 1410 | 91 | 1501 | 461 | 40 | 501 | 732 | 20 | 752 | 15375 | 1877 | 17252 | 10858 | 4700 | 15558 | 5886 | 1507 | 7393 | 294 | 86 | 380 |
| Anterior- and Retropatellar pain Incidence Rate (per 1000 person-years) |  |  |  |  |  |  |  |  |  |  |  |  |  |  |  |  |  |  |  |  |  |  |  |  |  |  |  |  |  |  |
| Army | 4.1 | ** | 4.1 | 5.1 | ** | 5.1 | 5.4 | ** | 5.4 | 6.1 | 9.2 | 6.2 | ** | ** | ** | 5.8 | 9.8 | 5.8 | 6.4 | 9.3 | 6.7 | 6.2 | 8.8 | 6.8 | 7.1 | 8.4 | 7.3 | 6.5 | 5.8 | 6.4 |
| Navy | 4.2 | ** | 4.2 | 2.7 | ** | 2.7 | 3.8 | ** | 3.8 | 2.3 | 4.1 | 2.5 | 2.8 | 5.3 | 3.0 | ** | ** | ** | 2.9 | 3.5 | 2.9 | 3.3 | 4.1 | 3.5 | 3.2 | 4.3 | 3.4 | 2.7 | 3.7 | 2.9 |
| Air Force | ** | ** | ** | ** | ** | ** | ** | ** | ** | ** | ** | ** | 6.7 | 5.7 | 6.6 | ** | ** | ** | 5.5 | 7.3 | 5.6 | 5.7 | 7.7 | 6.3 | 5.3 | 7.3 | 5.7 | ** | ** | ** |
| Marines | ** | ** | ** | 2.6 | ** | 2.6 | 2.5 | ** | 2.5 | 2.6 | ** | 2.6 | 2.7 | 3.7 | 2.7 | 2.2 | 3.8 | 2.2 | 3.7 | 5.4 | 3.8 | 3.6 | 5.7 | 3.9 | 3.5 | 4.6 | 3.6 | ** | ** | ** |
| Total | 4.1 | ** | 4.1 | 4.3 | ** | 4.3 | 4.7 | ** | 4.7 | 5.3 | 7.6 | 5.4 | 4.4 | 5.3 | 4.4 | 4.9 | 6.7 | 4.9 | 4.6 | 5.8 | 4.7 | 5.1 | 7.3 | 5.6 | 5.5 | 7.1 | 5.8 | 2.9 | 3.7 | 3.0 |
| Patellar Instability (n=11956) |  |  |  |  |  |  |  |  |  |  |  |  |  |  |  |  |  |  |  |  |  |  |  |  |  |  |  |  |  |  |
| Army | 35 | ** | 35 | 609 | ** | 609 | 77 | ** | 77 | 204 | 17 | 221 | ** | ** | 0 | 124 | 3 | 127 | 755 | 132 | 887 | 785 | 276 | 1061 | 525 | 169 | 694 | 8 | 0 | 8 |
| Navy | 7 | ** | 7 | 16 | ** | 16 | 0 | ** | 0 | 19 | 6 | 25 | 37 | 3 | 40 | ** | ** | ** | 838 | 191 | 1029 | 328 | 150 | 478 | 133 | 51 | 184 | 109 | 30 | 139 |
| Air Force | ** | ** | ** | ** | ** | ** | ** | ** | ** | 0 | 0 | ** | 50 | 4 | 54 | ** | ** | ** | 1014 | 139 | 1153 | 487 | 379 | 866 | 272 | 113 | 385 | ** | ** | ** |
| Marines | ** | ** | ** | 328 | ** | 328 | 18 | ** | 18 | 56 | 0 | 56 | 17 | 3 | 20 | 29 | 1 | 30 | 435 | 32 | 467 | 288 | 79 | 367 | 176 | 23 | 199 | ** | ** | ** |
| Total | 42 | ** | 42 | 953 | ** | 953 | 95 | ** | 95 | 279 | 23 | 302 | 104 | 10 | 114 | 153 | 4 | 157 | 3042 | 494 | 3536 | 1888 | 884 | 2772 | 1106 | 356 | 1462 | 117 | 30 | 147 |
| Patellar Instability Incidence Rate (per 1000 person-years) |  |  |  |  |  |  |  |  |  |  |  |  |  |  |  |  |  |  |  |  |  |  |  |  |  |  |  |  |  |  |
| Army | 0.6 | ** | 0.6 | 0.9 | ** | 0.9 | 0.9 | ** | 0.9 | 1.0 | 2.1 | 1.0 | ** | ** | ** | 1.1 | 2.1 | 1.1 | 0.9 | 1.6 | 0.9 | 0.9 | 1.1 | 0.9 | 1.1 | 1.6 | 1.2 | 1.9 | 0.0 | 1.7 |
| Navy | 0.5 | ** | 0.5 | 1.2 | ** | 1.2 | 0.0 | ** | 0.0 | 0.8 | 1.6 | 0.9 | 0.8 | 0.7 | 0.8 | ** | ** | ** | 0.7 | 1.2 | 0.8 | 0.8 | 1.2 | 0.9 | 0.9 | 1.4 | 1.0 | 1.1 | 1.3 | 1.1 |
| Air Force | ** | ** | ** | ** | ** | ** | ** | ** | ** | ** | ** | ** | 1.2 | 1.6 | 1.2 | ** | ** | ** | 1.1 | 2.2 | 1.2 | 1.0 | 1.7 | 1.2 | 1.1 | 2.1 | 1.3 | ** | ** | ** |
| Marines | ** | ** | ** | 1.1 | ** | 1.1 | 0.7 | ** | 0.7 | 1.6 | ** | 1.6 | 1.0 | 5.6 | 1.1 | 0.8 | 0.6 | 0.8 | 1.0 | 1.5 | 1.1 | 0.8 | 1.5 | 0.9 | 1.0 | 1.2 | 1.0 | ** | ** | ** |
| Total | 0.6 | ** | 0.6 | 1.0 | ** | 1.0 | 0.9 | ** | 0.9 | 1.1 | 1.9 | 1.1 | 1.0 | 1.3 | 1.0 | 1.0 | 1.3 | 1.0 | 0.9 | 1.5 | 1.0 | 0.9 | 1.4 | 1.0 | 1.0 | 1.7 | 1.1 | 1.1 | 1.3 | 1.2 |
| Knee Tendinopathy (n=78637) |  |  |  |  |  |  |  |  |  |  |  |  |  |  |  |  |  |  |  |  |  |  |  |  |  |  |  |  |  |  |
| Army | 169 | ** | 169 | 4050 | ** | 4050 | 534 | ** | 534 | 1726 | 90 | 1816 | ** | ** | 0 | 881 | 17 | 898 | 8186 | 1002 | 9188 | 7645 | 2539 | 10184 | 4466 | 1164 | 5630 | 54 | 11 | 65 |
| Navy | 30 | ** | 30 | 50 | ** | 50 | 5 | ** | 5 | 84 | 13 | 97 | 139 | 35 | 174 | ** | ** | ** | 3763 | 729 | 4492 | 1785 | 550 | 2335 | 706 | 179 | 885 | 394 | 155 | 549 |
| Air Force | ** | ** | ** | ** | ** | ** | ** | ** | ** | ** | 0 | ** | 234 | 27 | 261 | ** | ** | ** | 5860 | 468 | 6328 | 3705 | 1605 | 5310 | 1749 | 364 | 2113 | ** | ** | ** |
| Marines | ** | ** | ** | 966 | ** | 966 | 85 | ** | 85 | 99 | 0 | 99 | 45 | 1 | 46 | 129 | 7 | 136 | 2235 | 126 | 2361 | 2030 | 383 | 2413 | 1022 | 120 | 1142 | ** | ** | ** |
| Total | 199 | ** | 199 | 5066 | ** | 5066 | 624 | ** | 624 | 1909 | 103 | 2012 | 418 | 63 | 481 | 1010 | 24 | 1034 | 20044 | 2325 | 22369 | 15165 | 5077 | 20242 | 7943 | 1827 | 9770 | 448 | 166 | 614 |
| Knee Tendinopathy Incidence Rate (per 1000 person-years) |  |  |  |  |  |  |  |  |  |  |  |  |  |  |  |  |  |  |  |  |  |  |  |  |  |  |  |  |  |  |
| Army | 2.7 | ** | 2.7 | 6.2 | ** | 6.2 | 6.5 | ** | 6.5 | 8.4 | 11.0 | 8.5 | ** | ** | ** | 7.8 | 11.9 | 7.9 | 9.5 | 12.4 | 9.7 | 8.6 | 10.2 | 8.9 | 9.2 | 11.2 | 9.6 | 12.5 | 21.3 | 13.4 |
| Navy | 2.3 | ** | 2.3 | 3.7 | ** | 3.7 | 3.2 | ** | 3.2 | 3.4 | 3.4 | 3.4 | 3.2 | 7.7 | 3.6 | ** | ** | ** | 3.3 | 4.7 | 3.5 | 4.5 | 4.6 | 4.5 | 4.6 | 4.8 | 4.6 | 4.0 | 6.8 | 4.5 |
| Air Force | ** | ** | ** | ** | ** | ** | ** | ** | ** | ** | ** | ** | 5.4 | 11.0 | 5.7 | ** | ** | ** | 6.4 | 7.3 | 6.5 | 7.6 | 7.3 | 7.5 | 7.0 | 6.9 | 6.9 | ** | ** | ** |
| Marines | ** | ** | ** | 3.2 | ** | 3.2 | 3.4 | ** | 3.4 | 2.9 | ** | 2.9 | 2.5 | 1.9 | 2.5 | 3.5 | 4.5 | 3.5 | 5.3 | 5.8 | 5.4 | 6.0 | 7.1 | 6.1 | 5.7 | 6.4 | 5.7 | ** | ** | ** |
| Total | 2.7 | ** | 2.7 | 5.2 | ** | 5.2 | 5.7 | ** | 5.7 | 7.2 | 8.5 | 7.2 | 4.0 | 8.4 | 4.3 | 6.7 | 8.0 | 6.8 | 6.0 | 7.2 | 6.1 | 7.2 | 7.9 | 7.3 | 7.4 | 8.6 | 7.6 | 4.4 | 7.2 | 4.9 |
| Army | 454 | ** | 454 | 7967 | ** | 7967 | 1054 | ** | 1054 | 3194 | 182 | 3376 | ** | ** | 0 | 1656 | 34 | 1690 | 14501 | 1892 | 16393 | 13957 | 5019 | 18976 | 8414 | 2206 | 10620 | 90 | 14 | 104 |
| Navy | 92 | ** | 92 | 102 | ** | 102 | 11 | ** | 11 | 160 | 35 | 195 | 299 | 62 | 361 | ** | ** | ** | 7874 | 1455 | 9329 | 3431 | 1195 | 4626 | 1330 | 389 | 1719 | 769 | 268 | 1037 |
| Air Force | ** | ** | ** | ** | ** | ** | ** | ** | ** | ** | ** | ** | 574 | 45 | 619 | ** | ** | ** | 11848 | 1074 | 12922 | 6967 | 3678 | 10645 | 3352 | 866 | 4218 | ** | ** | ** |
| Marines | ** | ** | ** | 2076 | ** | 2076 | 166 | ** | 166 | 244 | ** | 244 | 110 | 6 | 116 | 239 | 14 | 253 | 4238 | 275 | 4513 | 3556 | 769 | 4325 | 1839 | 229 | 2068 | ** | ** | ** |
| Total | 546 | ** | 546 | 10145 | ** | 10145 | 1231 | ** | 1231 | 3598 | 217 | 3815 | 983 | 113 | 1096 | 1895 | 48 | 1943 | 38461 | 4696 | 43157 | 27911 | 10661 | 38572 | 14935 | 3690 | 18625 | 859 | 282 | 1141 |
| Total Anterior Knee Pain Incidence Rate (per 1000 person-years) |  |  |  |  |  |  |  |  |  |  |  |  |  |  |  |  |  |  |  |  |  |  |  |  |  |  |  |  |  |  |
| Army | 7.4 | ** | 7.4 | 12.3 | ** | 12.3 | 12.8 | ** | 12.8 | 15.5 | 22.2 | 15.8 | ** | ** | ** | 14.7 | 23.8 | 14.8 | 16.8 | 23.3 | 17.3 | 15.7 | 20.1 | 16.7 | 17.4 | 21.3 | 18.1 | 20.8 | 27.0 | 21.5 |
| Navy | 6.9 | ** | 6.9 | 7.6 | ** | 7.6 | 7.0 | ** | 7.0 | 6.4 | 9.1 | 6.8 | 6.8 | 13.7 | 7.5 | ** | ** | ** | 6.9 | 9.5 | 7.2 | 8.6 | 9.9 | 8.9 | 8.6 | 10.5 | 9.0 | 7.8 | 11.8 | 8.6 |
| Air Force | ** | ** | ** | ** | ** | ** | ** | ** | ** | ** | ** | ** | 13.2 | 18.3 | 13.5 | ** | ** | ** | 13.0 | 16.7 | 13.3 | 14.3 | 16.7 | 15.0 | 13.3 | 16.3 | 13.9 | ** | ** | ** |
| Marines | ** | ** | ** | 6.8 | ** | 6.8 | 6.7 | ** | 6.7 | 7.0 | ** | 7.0 | 6.2 | 11.2 | 6.3 | 6.4 | 9.0 | 6.5 | 10.1 | 12.6 | 10.3 | 10.5 | 14.3 | 11.0 | 10.2 | 12.3 | 10.4 | ** | ** | ** |
| Total | 7.3 | ** | 7.3 | 10.5 | ** | 10.5 | 11.3 | ** | 11.3 | 13.5 | 18.0 | 13.7 | 9.3 | 15.0 | 9.7 | 12.6 | 16.1 | 12.7 | 11.5 | 14.6 | 11.8 | 13.2 | 16.5 | 14.0 | 14.0 | 17.3 | 14.5 | 8.4 | 12.1 | 9.1 |

**Appendix B.** Numbers and incidence of anterior knee pain among officers in the US Armed Forces across occupation.

|  | Ground and Naval<br>Gunfire |  |  | Aviation |  |  | Engineering &<br>Maintenance |  |  | Administration |  |  | Operations &<br>Intelligence |  |  | Logistics |  |  | Services |  |  |
| --- | --- | --- | --- | --- | --- | --- | --- | --- | --- | --- | --- | --- | --- | --- | --- | --- | --- | --- | --- | --- | --- |
|  | M | F | T | M | F | T | M | F | T | M | F | T | M | F | T | M | F | T | M | F | T |
| Anterior- and Retropatellar pain (n=10105) |  |  |  |  |  |  |  |  |  |  |  |  |  |  |  |  |  |  |  |  |  |
| Army | 909 | 11 | 920 | 354 | 24 | 378 | 735 | 144 | 879 | 346 | 189 | 535 | 385 | 94 | 479 | 478 | 192 | 670 | 696 | 384 | 1080 |
| Navy | 280 | 38 | 318 | 214 | 16 | 230 | 204 | 21 | 225 | 94 | 29 | 123 | 104 | 23 | 127 | 93 | 23 | 116 | 241 | 150 | 391 |
| Air Force | 27 | 1 | 28 | 494 | 48 | 542 | 296 | 64 | 360 | 150 | 88 | 238 | 339 | 98 | 437 | 227 | 68 | 295 | 386 | 328 | 714 |
| Marines | 111 | 5 | 116 | 78 | 3 | 81 | 93 | 6 | 99 | 107 | 21 | 128 | 56 | 6 | 62 | 87 | 14 | 101 | 18 | 5 | 23 |
| Total | 1327 | 55 | 1382 | 1140 | 91 | 1231 | 1328 | 235 | 1563 | 697 | 327 | 1024 | 884 | 221 | 1105 | 885 | 297 | 1182 | 1341 | 867 | 2208 |
| Anterior- and Retropatellar pain Incidence Rate (per 1000 person-years) |  |  |  |  |  |  |  |  |  |  |  |  |  |  |  |  |  |  |  |  |  |
| Army | 4.7 | 6.3 | 4.7 | 4.1 | 5.0 | 4.1 | 5.9 | 7.1 | 6.1 | 6.5 | 9.5 | 7.3 | 5.4 | 7.0 | 5.7 | 6.8 | 9.6 | 7.4 | 4.5 | 5.9 | 4.9 |
| Navy | 3.2 | 3.8 | 3.3 | 2.2 | 2.7 | 2.2 | 3.6 | 6.5 | 3.8 | 4.6 | 5.3 | 4.8 | 3.6 | 5.0 | 3.8 | 4.2 | 6.6 | 4.5 | 3.0 | 3.6 | 3.2 |
| Air Force | 4.9 | 1.4 | 4.5 | 3.1 | 5.8 | 3.2 | 3.9 | 5.4 | 4.1 | 4.5 | 6.6 | 5.1 | 4.5 | 5.7 | 4.7 | 5.8 | 6.7 | 6.0 | 3.9 | 5.8 | 4.6 |
| Marines | 2.8 | 11.7 | 2.9 | 1.8 | 2.4 | 1.9 | 4.2 | 5.4 | 4.3 | 5.4 | 7.2 | 5.6 | 3.0 | 4.8 | 3.1 | 3.5 | 4.6 | 3.7 | 3.6 | 7.2 | 4.0 |
| Total | 4.1 | 4.3 | 4.1 | 3.0 | 4.5 | 3.0 | 4.8 | 6.4 | 5.0 | 5.5 | 7.9 | 6.1 | 4.6 | 6.0 | 4.8 | 5.7 | 8.1 | 6.1 | 3.9 | 5.3 | 4.4 |
| Patellar Instability (n=1014) |  |  |  |  |  |  |  |  |  |  |  |  |  |  |  |  |  |  |  |  |  |
| Army | 76 | 1 | 77 | 27 | 1 | 28 | 53 | 17 | 70 | 20 | 16 | 36 | 27 | 13 | 40 | 15 | 14 | 29 | 61 | 40 | 101 |
| Navy | 27 | 7 | 34 | 26 | 1 | 27 | 22 | 3 | 25 | 2 | 2 | 4 | 11 | 1 | 12 | 13 | 3 | 16 | 19 | 26 | 45 |
| Air Force | 6 | 1 | 7 | 63 | 4 | 67 | 33 | 15 | 48 | 15 | 10 | 25 | 35 | 22 | 57 | 23 | 12 | 35 | 50 | 44 | 94 |
| Marines | 10 | 0 | 10 | 15 | 0 | 15 | 8 | 2 | 10 | 5 | 3 | 8 | 5 | 0 | 5 | 13 | 1 | 14 | 0 | 0 | 0 |
| Total | 119 | 9 | 128 | 131 | 6 | 137 | 116 | 37 | 153 | 42 | 31 | 73 | 78 | 36 | 114 | 64 | 30 | 94 | 130 | 110 | 240 |
| Patellar Instability Incidence Rate (per 1000 person-years) |  |  |  |  |  |  |  |  |  |  |  |  |  |  |  |  |  |  |  |  |  |
| Army | 0.4 | 0.6 | 0.4 | 0.3 | 0.2 | 0.3 | 0.4 | 0.8 | 0.5 | 0.4 | 0.8 | 0.5 | 0.4 | 1.0 | 0.5 | 0.2 | 0.7 | 0.3 | 0.4 | 0.6 | 0.5 |
| Navy | 0.3 | 0.7 | 0.3 | 0.3 | 0.2 | 0.3 | 0.4 | 0.9 | 0.4 | 0.1 | 0.4 | 0.2 | 0.4 | 0.2 | 0.4 | 0.6 | 0.9 | 0.6 | 0.2 | 0.6 | 0.4 |
| Air Force | 1.1 | 1.4 | 1.1 | 0.4 | 0.5 | 0.4 | 0.4 | 1.3 | 0.5 | 0.5 | 0.8 | 0.5 | 0.5 | 1.3 | 0.6 | 0.6 | 1.2 | 0.7 | 0.5 | 0.8 | 0.6 |
| Marines | 0.3 | 0.0 | 0.3 | 0.4 | 0.0 | 0.3 | 0.4 | 1.8 | 0.4 | 0.3 | 1.0 | 0.3 | 0.3 | 0.0 | 0.2 | 0.5 | 0.3 | 0.5 | 0.0 | 0.0 | 0.0 |
| Total | 0.4 | 0.7 | 0.4 | 0.3 | 0.3 | 0.3 | 0.4 | 1.0 | 0.5 | 0.3 | 0.7 | 0.4 | 0.4 | 1.0 | 0.5 | 0.4 | 0.8 | 0.5 | 0.4 | 0.7 | 0.5 |
| Knee Tendinopathy (n=3216) |  |  |  |  |  |  |  |  |  |  |  |  |  |  |  |  |  |  |  |  |  |
| Army | 864 | 8 | 872 | 208 | 10 | 218 | 647 | 126 | 773 | 277 | 109 | 386 | 368 | 70 | 438 | 404 | 125 | 529 | 573 | 247 | 820 |
| Navy | 246 | 28 | 274 | 187 | 11 | 198 | 184 | 8 | 192 | 77 | 25 | 102 | 100 | 21 | 121 | 80 | 17 | 97 | 183 | 128 | 311 |
| Air Force | 25 | 2 | 27 | 393 | 27 | 420 | 373 | 53 | 426 | 144 | 71 | 215 | 310 | 65 | 375 | 205 | 58 | 263 | 370 | 209 | 579 |
| Marines | 110 | 1 | 111 | 85 | 2 | 87 | 97 | 3 | 100 | 73 | 17 | 90 | 67 | 5 | 72 | 89 | 9 | 98 | 26 | 1 | 27 |
| Total | 1245 | 39 | 1284 | 873 | 50 | 923 | 1301 | 190 | 1491 | 571 | 222 | 793 | 845 | 161 | 1006 | 778 | 209 | 987 | 1152 | 585 | 1737 |
| Knee Tendinopathy Incidence Rate (per 1000 person-years) |  |  |  |  |  |  |  |  |  |  |  |  |  |  |  |  |  |  |  |  |  |
| Army | 4.5 | 4.6 | 4.5 | 2.4 | 2.1 | 2.4 | 5.2 | 6.2 | 5.4 | 5.2 | 5.5 | 5.3 | 5.2 | 5.2 | 5.2 | 5.8 | 6.2 | 5.9 | 3.7 | 3.8 | 3.7 |
| Navy | 2.8 | 2.8 | 2.8 | 1.9 | 1.9 | 1.9 | 3.3 | 2.5 | 3.2 | 3.8 | 4.5 | 3.9 | 3.5 | 4.6 | 3.7 | 3.6 | 4.9 | 3.8 | 2.2 | 3.0 | 2.5 |
| Air Force | 4.6 | 2.8 | 4.4 | 2.5 | 3.3 | 2.5 | 4.9 | 4.5 | 4.8 | 4.3 | 5.4 | 4.6 | 4.1 | 3.8 | 4.0 | 5.3 | 5.7 | 5.4 | 3.8 | 3.7 | 3.8 |
| Marines | 2.8 | 2.3 | 2.8 | 2.0 | 1.6 | 2.0 | 4.4 | 2.7 | 4.3 | 3.7 | 5.8 | 3.9 | 3.6 | 4.0 | 3.6 | 3.6 | 3.0 | 3.5 | 5.2 | 1.4 | 4.7 |
| Total | 3.8 | 3.0 | 3.8 | 2.3 | 2.5 | 2.3 | 4.7 | 5.2 | 4.7 | 4.5 | 5.3 | 4.7 | 4.4 | 4.4 | 4.4 | 5.0 | 5.7 | 5.1 | 3.4 | 3.6 | 3.4 |
| Total Anterior Knee Pain (n=14335) |  |  |  |  |  |  |  |  |  |  |  |  |  |  |  |  |  |  |  |  |  |
| Army | 1849 | 20 | 1869 | 589 | 35 | 624 | 1435 | 287 | 1722 | 643 | 314 | 957 | 780 | 177 | 957 | 897 | 331 | 1228 | 1330 | 671 | 2001 |
| Navy | 553 | 73 | 626 | 427 | 28 | 455 | 410 | 32 | 442 | 173 | 56 | 229 | 215 | 45 | 260 | 186 | 43 | 229 | 443 | 304 | 747 |
| Air Force | 58 | 4 | 62 | 950 | 79 | 1029 | 702 | 132 | 834 | 309 | 169 | 478 | 684 | 185 | 869 | 455 | 138 | 593 | 806 | 581 | 1387 |
| Marines | 231 | 6 | 237 | 178 | 5 | 183 | 198 | 11 | 209 | 185 | 41 | 226 | 128 | 11 | 139 | 189 | 24 | 213 | 44 | 6 | 50 |
| Total | 2691 | 103 | 2794 | 2144 | 147 | 2291 | 2745 | 462 | 3207 | 1310 | 580 | 1890 | 1807 | 418 | 2225 | 1727 | 536 | 2263 | 2623 | 1562 | 4185 |
| Total Anterior Knee Pain Incidence Rate (per 1000 person-years) |  |  |  |  |  |  |  |  |  |  |  |  |  |  |  |  |  |  |  |  |  |
| Army | 9.6 | 11.4 | 9.6 | 6.8 | 7.3 | 6.8 | 11.6 | 14.1 | 11.9 | 12.0 | 15.8 | 13.0 | 11.0 | 13.1 | 11.4 | 12.8 | 16.5 | 13.6 | 8.5 | 10.3 | 9.1 |
| Navy | 6.3 | 7.3 | 6.4 | 4.4 | 4.7 | 4.4 | 7.3 | 10.0 | 7.5 | 8.5 | 10.1 | 8.9 | 7.5 | 9.8 | 7.9 | 8.4 | 12.3 | 8.9 | 5.4 | 7.2 | 6.0 |
| Air Force | 10.6 | 5.7 | 10.0 | 6.0 | 9.6 | 6.2 | 9.2 | 11.1 | 9.5 | 9.3 | 12.8 | 10.3 | 9.1 | 10.8 | 9.4 | 11.7 | 13.6 | 12.1 | 8.2 | 10.3 | 9.0 |
| Marines | 5.9 | 14.1 | 5.9 | 4.2 | 4.0 | 4.2 | 9.0 | 9.9 | 9.1 | 9.3 | 14.0 | 9.9 | 6.8 | 8.8 | 6.9 | 7.7 | 7.9 | 7.7 | 8.8 | 8.7 | 8.8 |
| Total | 8.3 | 8.0 | 8.3 | 5.6 | 7.3 | 5.7 | 9.9 | 12.6 | 10.2 | 10.3 | 13.9 | 11.2 | 9.3 | 11.4 | 9.7 | 11.1 | 14.6 | 11.8 | 7.7 | 9.5 | 8.3 |

## Notes

### Competing Interest Statement

The authors have declared no competing interest.

### Funding Statement

The authors are military service members or employees of the U.S. Government. This work was prepared as part of their official duties.

### Author Declarations

The study protocol was approved by the Naval Health Research Center Institutional Review Board in compliance with all applicable Federal regulations governing the protection of human subjects. Research data were derived from an approved Naval Health Research Center Institutional Review Board protocol, number NHRC.2020.0203-NHSR.

